# Lack of the human choline transporter-like protein CTL2 causes hearing impairment and a rare red blood cell phenotype

**DOI:** 10.1101/2022.05.13.22273920

**Authors:** Bérengère Koehl, Cédric Vrignaud, Mahmoud Mikdar, Thankam S. Nair, Lucy Yang, Guy Laiguillon, Sophie Anselme-Martin, Claudine Giroux-Lathuile, Hanane El Kenz, Olivier Hermine, Narla Mohandas, Jean Pierre Cartron, Yves Colin, Olivier Detante, Caroline Le Van Kim, Thomas E. Carey, Slim Azouzi, Thierry Peyrard

**Author notes:** To whom correspondence should be addressed: Slim. Azouzi, Inserm, UMR_S1134, INTS, 6 rue Alexandre Cabanel, 75015 Paris, France., Thierry Peyrard, Inserm, UMR_S1134, INTS, 6 rue Alexandre Cabanel, 75015 Paris, France. These authors contributed equally to this work (Bérengère Koehl, Cédric Vrignaud, Slim Azouzi, Thierry Peyrard).

## Abstract

Recent genome-wide association and murine studies identified the human neutrophil antigen -3a/b polymorphism (HNA-3a/b) in *SLC44A2* (rs2288904-G/A) as a risk factor in venous thromboembolism (VTE). The choline transporter-like protein CTL2 encoded by the *SLC44A2* gene plays an important role in platelet aggregation and neutrophil interaction with the von Willebrand factor. By investigating alloantibodies to a high-prevalence antigen of unknown specificity, found in patients with a rare blood type, we showed that CTL2 is also expressed in red blood cells and carries a new blood group system. Furthermore, we identified three siblings of European ancestry who are homozygous for a large deletion in *SLC44A2*, resulting in complete CTL2 deficiency. Interestingly, the first-ever reported CTL2-deficient individuals suffer from progressive hearing impairment, recurrent arterial aneurysms and epilepsy. In contrast to *Slc44a2*^-/-^ mice, CTL2_null_ individuals showed normal platelet aggregation and do not suffer from any apparent hematological disorders. In addition, CD34^+^ cells from CTL2_null_ patients undergo normal *ex vivo* erythropoiesis, indicating that CTL2 is not essential for erythroid proliferation and differentiation. Overall, our findings confirm the function of CTL2 in hearing preservation and provide new insights into the possible role of this protein in maintaining cerebrovascular homeostasis.

## Introduction

Blood group antigens are defined by the presence or absence of specific antigens on the surface of the red blood cell (RBC) membrane and are inherited characteristics resulting from genetic polymorphism at the identified blood group loci^1^. A null RBC phenotype, in which all antigens in one system are absent, has been identified in most blood group systems, generally because of the complete absence of the antigen carrier molecule from the RBC membrane. In many cases, individuals with these null phenotypes are apparently healthy, suggesting that the biological function of the missing protein may be compensated by another mechanism^2-5^. However, in some cases, the null phenotype is associated with mild to severe hematological and/or non-hematological disorders^6^. Therefore, null phenotypes represent natural ‘knockouts’ and represent unique opportunities in providing indications towards the function of membrane proteins, not only in erythroid cells but also in other cells or tissues^7-12^.

SLC44A2 is a member of the choline transporter-like (CTL) family and is present in various human tissues including kidney, lung, inner ear, endothelial and blood cells. Currently, the molecular and cellular function of SLC44A2 is not well-defined. In murine models, its deficiency has been associated with hearing loss, altered neutrophil recruitment and impaired platelet activation^13,14^. In humans, *SLC44A2* has been identified as a susceptibility locus for venous thromboembolism (VTE), which generated heightened interest in its function and attention has shifted to its role in thrombosis^15,16^. Several studies have recently confirmed that Slc44a2 promotes thrombosis in a mouse model of laser injury and venous stenosis and suggest that this may be related to platelet-neutrophil interaction^17^. Accordingly, a recent study identified activated α_IIb_β_3_ integrin in platelets as a receptor and agonist for neutrophils through SLC44A2^18^. In addition, a murine study showed that Slc44a2 is a mitochondrial choline transporter that regulates mitochondrial synthesis of ATP and platelet activation^13^. The mitochondrial function of this protein was also reported in the human brain microvascular endothelial cells of the blood-brain barrier. It has been proposed that SLC44A2 is responsible for choline transport into mitochondria, an important step in the oxidative pathway of choline metabolism^19,20^.

Herein, we show that SLC44A2 is expressed in erythrocytes and carries new blood group antigens called RIF and VER, establishing CTL2 as the 39^th^ blood group system recognized by the ISBT. Interestingly, we demonstrate that a large homozygous deletion in *SLC44A2* gene results in the total absence of this protein in three siblings, which all suffer from age-related hearing loss, and some of them from epilepsy or intracranial aneurysms. Unexpectedly, the absence of SLC44A2 from platelets and RBCs does not cause any apparent hematological disorder. Finally, we demonstrated that those two new anti-SLC44A2 red cell alloantibodies lead to neutrophil activation and their adhesion on endothelial cells.

## Results

### A single nucleotide variation in the *SLC44A2* gene encodes a new blood group antigen

As part of an effort to identify the genes encoding high-prevalence blood group antigens with an unknown molecular basis, we focused here our research on the serum of a pregnant woman of North African ancestry who developed an RBC antibody during her pregnancy. A multilaboratory investigation was initially inconclusive, and the antibody was claimed to be of unknown specificity. We propose calling this antibody anti-RIF and the new high-prevalence antigen RIF. Further testing of this antibody with several RBC samples lacking a high-prevalence antigen of unknown specificity allowed the identification of six more RIF– samples from five North African and one individual of European descent. To identify the gene underlying the RIF antigen, we performed whole-exome sequencing (WES) of these 7 RIF– individuals. After analysis of WES data, three shared variants (with minor allele frequency, MAF < 1%) were found in the six RIF– North African individuals but not in the European proband: one in *SLC44A2* (c.1192C>A [p.Pro398Thr] [GenBank: NM_001145056]), a second in *PPAN-P2RY11* (c.822+31T>C) [GenBank: NM_020230]), and a third in *TYK2* (c.2036G>C. [p.Arg679Pro] [GenBank: NM_003331]) genes. These three genes are located on chromosome 19p13.2 (Figure 1A). The *SLC44A2, PPAN-P2RY11* and *TYK2* variants are all present in the Genome Aggregation Database (gnomAD) browser, but the MAF values are available only for *PPAN-P2RY11* (0.00003) and *TYK2* (0.0002). *SLC44A2* was considered the most prominent candidate because this gene encodes the choline transporter-like 2 (CTL2), a membrane protein expressed in blood cells^21-24^. Family segregation confirmed that proband 1 (RIF–) was homozygous for these variants, while her children and sibling were heterozygous for the *SLC44A2* variant (RIF-/RIF+ phenotype) (Figure 1B). To determine if the *SLC44A2* polymorphism is responsible for the RIF– blood type, we transfected murine L-929 cells with an *SLC44A2* expression vector and analyzed them by flow cytometry using the anti-RIF anti-sera. Exogenous expression of *SLC44A2* in L-929 cells was responsible for cell surface expression of the RIF antigen. In contrast, the anti-RIF antibody failed to bind the L-929 cells expressing the p.Pro398Thr mutant of *SLC44A2* encoded by the c.1192C>A variant found in the RIF– proband 1 (Figure 1C). Altogether, these results demonstrated that the RIF– phenotype results from the p.Pro398Thr substitution in CTL2. This finding allowed us to develop a genotyping assay to identify rare RIF– blood donors. An AS-PCR assay for the c.1192C>A variant (Figure 1D) confirmed the homozygous and heterozygous state of the *SLC44A2* mutation in the proband and her sibling, respectively, in accordance with their RIF phenotype (Supplemental Figure 1).

**Figure 1:**
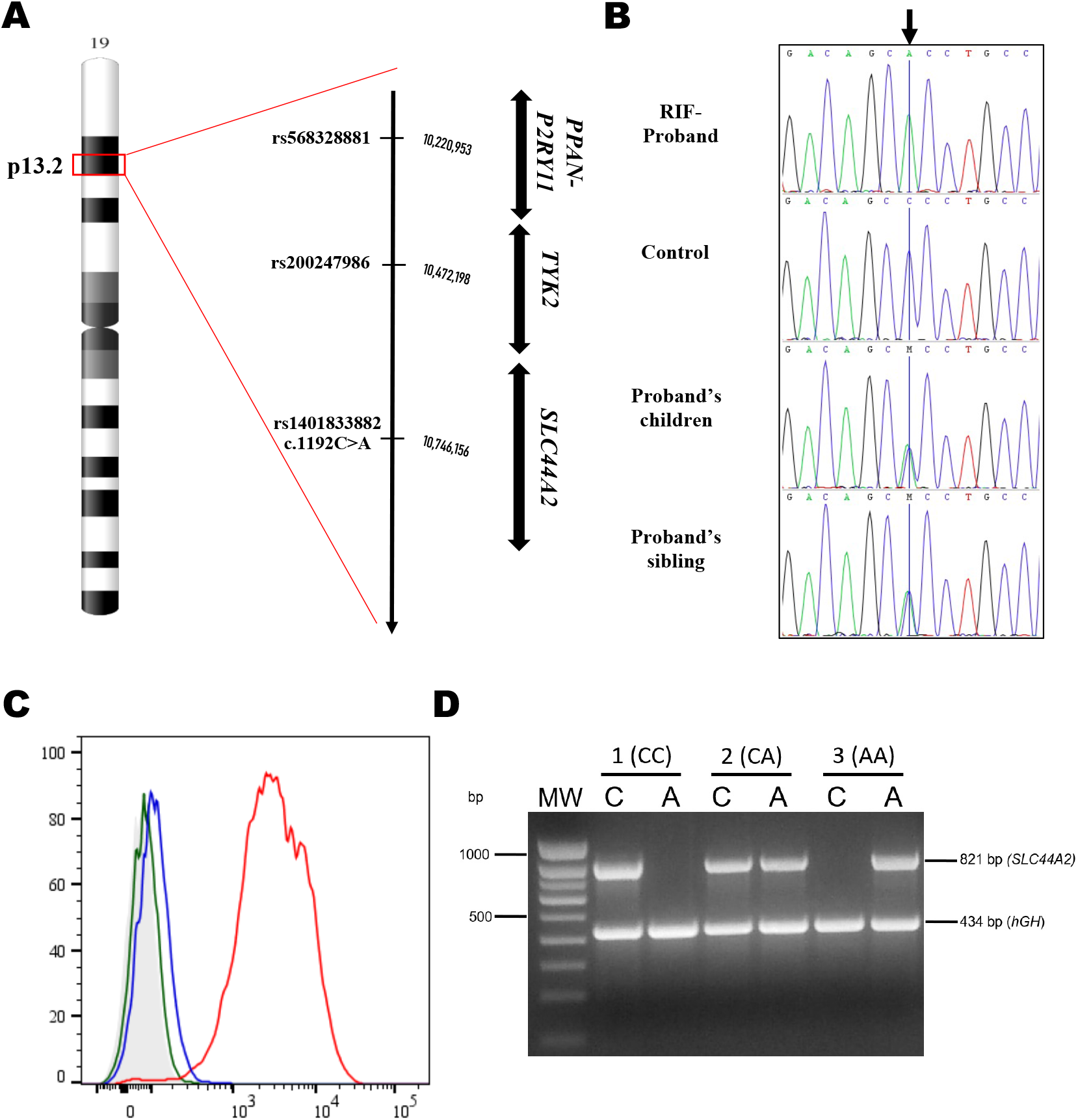
Molecular basis of the RIF blood group antigen. **A**. Common variant candidates found in RIF– North African individuals were shown in an ideogram of chromosome 19, highlighting a region of 19p13.2 with a red box. An expanded view of this region is shown, including the genes and chromosomal coordinates. **B**. Sequencing chromatograms of the *SLC44A2* variant in proband 1’s family. Proband 1’s child and sibling are heterozygous, whereas the RIF– proband is homozygous for the c.1192C>A mutation (noted by the black arrow). **C**. Exogenous expression of SLC44A2 in L-929 cells results in cell surface expression of the RIF antigen. L-929 cells stably transfected with pcDNA3.1/SLC44A2 full-length cDNA (red) or with cDNA3.1/SLC44A2(1192A) (blue) and then analyzed by flow cytometry with anti-RIF antibody. The green profile corresponds to untransfected L-929 (wild type), and the light gray profile corresponds to wild-type L-929 cells incubated with only the secondary antibody. **D**. Representative genotyping of the c.1192C>A mutation by allele-specific PCR. For each sample, two PCRs were performed, one with a specific primer for the 1192C allele (PCR 1) and a second with specific primers for the 1192A allele (PCR 2). An 821-bp band indicated the presence of the allele; amplification failure indicated the absence of the allele. The 434 bp amplification product of the *hGH* control primer is presented in all lanes. In the gel, the first lane (MW) shows the 100 bp DNA ladder marker (Eurogentec, Belgium); for the other lanes, C and A indicate the product from PCR 1 and PCR 2, respectively, amplified from RIF^+/+^ (1(CC)), RIF^+/-^ (2(CA)) and RIF^-/-^ (3(AA)) samples.

### A large deletion in the *SLC44A2* gene underlies a null phenotype

WES data from the RIF– European proband 2 indicated the absence of the c.1192C>A mutation in the *SLC44A2* gene despite her RBCs not reacting with anti-RIF antibody (Figure 2A). To solve this discrepancy, we first performed additional serological investigation by testing her RBC antibody onto the RBCs of a RIF– North African individual. Flow cytometry experiments showed that this alloantibody was not an anti-RIF because it reacted with RIF– RBCs (Figure 2B). We propose to call this antibody anti-VER. Furthermore, the fact that this antibody was developed by a RIF– proband but was not an anti-RIF suggests that this European proband does not exhibit the same molecular basis as the North African individuals. Next, we checked the coverage of the 22 exons of *SLC44A2* by analyzing copy-number variation (CNV) data and identified a putative homozygous intragenic deletion in *SLC44A2*, which completely removed the first fifteen exons along with the 5’ UTR region (Figure 2C). Using a primer walking approach and Sanger sequencing, the precise breakpoints of the deletion in the intergenic region and intron 14 were identified, with coordinates from chromosome 19:10,598,733-10,636,021 (GRCh38/hg38 assembly) (Figure 2C). This deletion defines a null allele of *SLC44A2*, explaining why the European proband had developed an alloantibody to SLC44A2/CTL2 (anti-VER). Furthermore, flow cytometry analysis of L-929 cells stably transfected with the *SLC44A2* cDNA showed cell surface expression of the VER antigen, confirming that the anti-VER antibody is directed against the SLC44A2 protein (Figure 3A). We then tested the binding of antibodies from RIF– individuals as well as from SLC44A2_null_ proband 2 (VER–) on recombinant human SLC44A2 protein (rHuSLC44A2) produced in transfected Sf9 insect cells^25^. rHuSLC44A2, produced in Sf9 cells either from whole cell lysates or immunoprecipitated with rabbit anti SLC44A2 (CTL2-NT)^26,27^, was used to assess serum alloantibodies from probands 1 and 2. As shown in Figure 3B, both RIF and VER antibodies reacted specifically with whole cell lysates or with rHuSLC44A2 concentrated by immunoprecipitation. Serum from proband 1 exhibited a higher titer and required only a 30 second exposure of the ECL blot to show binding to the rHuSLC44A2 protein in Sf9 whole cell lysates (WCL) (Figure 3B, lane 1), whereas the proband 2 serum required a 10-minute exposure to reach the same saturation (Figure 3B, lane 2). In contrast, when the rHuSLC44A2 protein was concentrated by immunoprecipitation, binding was detectable in 30 seconds with both anti-VER and anti-RIF sera (Figure 3B, lanes 3 and 4). Both sera also reacted against immunoprecipitated SLC44A2 from a human squamous cancer cell line (UM-SCC-47) that expresses high levels of the protein (Figure 3C, lanes 1 and 2). In another approach, the alloantibodies from proband 1 and 2 were also tested against cell extracts of wild-type and *Slc44a2* knockout mice ^28^. The VER serum, which arose in SLC44A2_null_ proband 2, binds well to the wild-type mouse lung Slc44a2 protein but not to the knockout murine lung protein, supporting the specificity of the alloantibodies for SLC44A2. A review of the mouse amino acid sequences showed that most of the murine Slc44A2 protein is homologous to the human form, but the region carrying the RIF antigen in humans was dissimilar in the mouse. The mouse lacks c.1192proline and has alanine in that site, explaining why anti-RIF fails to bind to the wild-type mouse protein (Figure 3D, right panel) (amino acid sequence comparison in Supplemental Figure 2).

**Figure 2:**
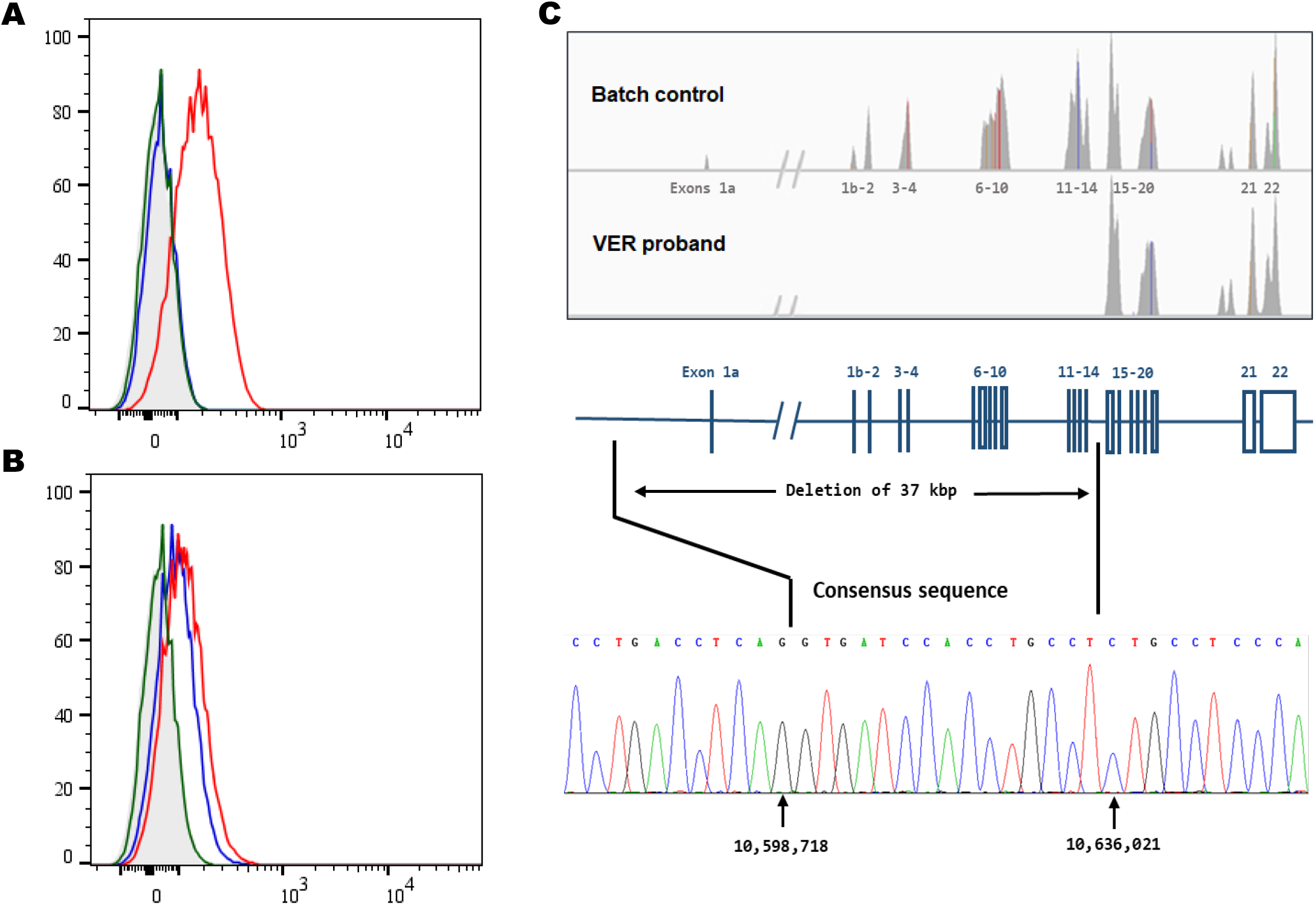
Null allele of *SLC44A2* is responsible for the rare VER– blood type. **A and B**. Flow analysis of SLC44A2 expression at the surface of RBCs from control donors (red), RIF– proband 1 (blue), and VER– proband 2 (green) using anti-RIF (**A**) and anti-VER antibodies (**B**). The light gray profile corresponds to RBCs incubated with only the secondary antibody. **C**. Integrative Genomics Viewer analysis reveals homozygous deletion of exons from 1 to 14 of the *SLC44A2* gene in VER– proband. Schematic representation of the *SLC44A2* gene and Sanger sequencing confirmation of the deletion breakpoints in genomic DNA of VER– proband.

**Figure 3:**
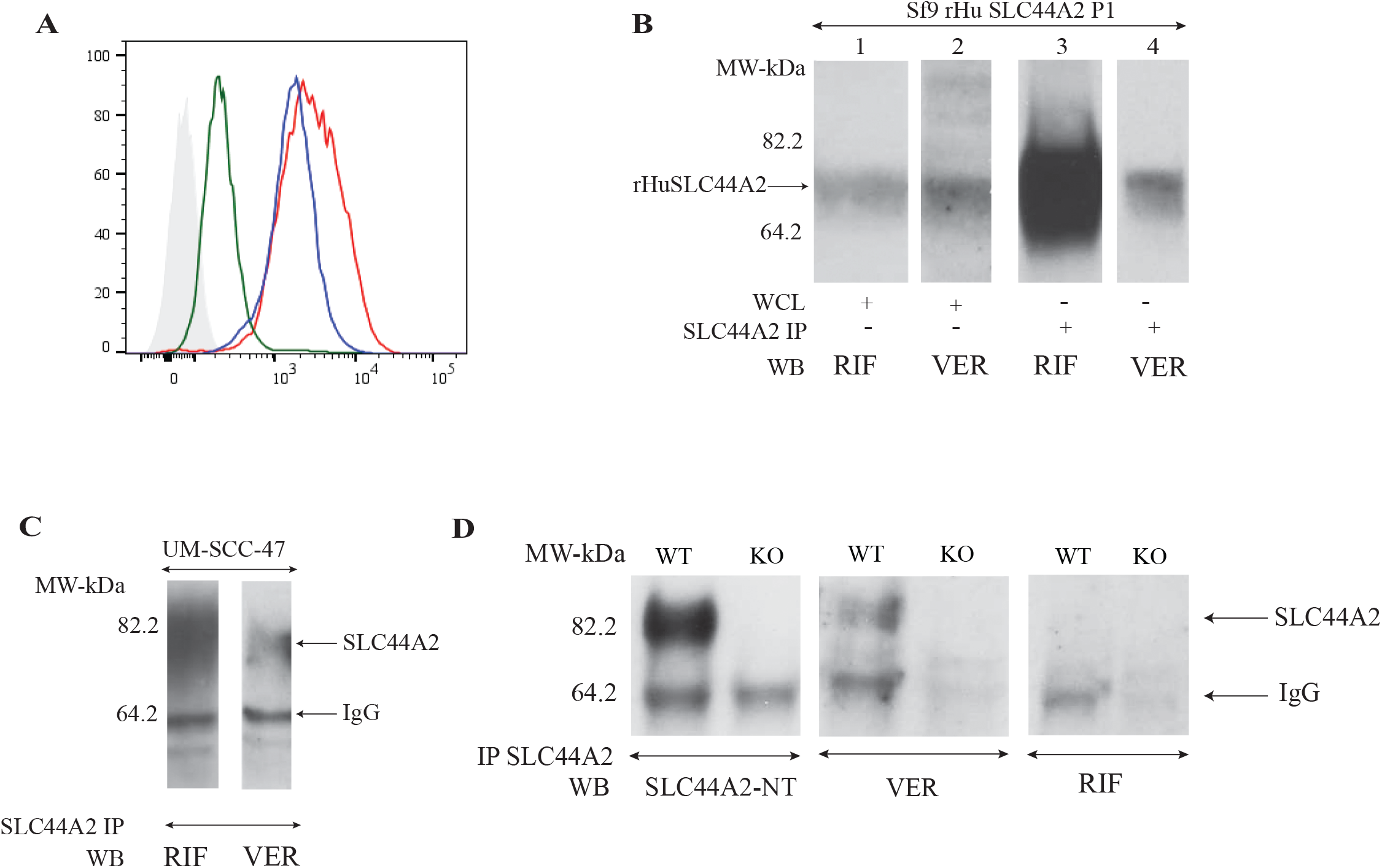
Reactivity of VER and RIF antibodies. **A**. Exogenous expression of SLC44A2 in L-929 cells results in cell surface expression of the VER antigen. L-929 cells stably transfected with pcDNA3.1/SLC44A2 full-length cDNA (red) or with cDNA3.1/SLC44A2(1192A) (blue) and then analyzed by flow cytometry with anti-VER antibody. The green profile corresponds to untransfected L-929 (wild type), and the light gray profile corresponds to wild-type cells incubated with only the secondary antibody. **B**. Western blotting shows that the anti-RIF and anti-VER antibodies bind to HuSLC44A2. Western blots of whole Sf9 lysates (WCL) expressing full-length recombinant human *SLC44A2* cDNA (rHuSLC44A2) were probed with RIF and VER antisera (1:10). Lanes 1 and 2 show strong binding to rHuSLC44A2. Titer differences in the sera required longer exposure (10 min) of the filter to the film to develop the image with VER serum compared to only 30 second exposure for RIF serum. A similar blot of immunoprecipitated rHuSLC44A2 from the Sf9 lysates similarly reacted with RIF and VER sera (lanes 3 and 4) showed equivalent exposure times (30 seconds for both sera) when tested against the concentrated rHuSLC44A2 protein. **C**. The anti RIF and VER antibodies bound to immunoprecipitated wild-type SLC44A2 in lysates of the UM-SCC-47 human squamous cancer cell line (exposure time 5 min). **D**. Western blot of murine Slc44a2 protein immunoprecipitated from wild-type and *Slc44a2* knockout mice with rabbit anti CTL2-NT serum was probed with anti CTL2-NT (left panel) and with VER serum (center Panel) or RIF serum (right panel). The left panel was developed with goat anti rabbit IgG heavy and light chain (Bethyl Lab cat # A120-113P); the center and right panels were developed with rabbit anti human IgG and IgM heavy and light chain specific (Jackson Immunochemicals cat # 309035107).

### Null allele of *SLC44A2* is associated with hearing loss

Proband 2, a European in their 60s homozygous for a null *SLC44A2* allele, was the first ever-reported individual with no expression of CTL2. Clinically, she suffered from major and unusual cerebrovascular anomalies that started in her 30s. She showed a giant intracranial aneurysm (ICA) of the right posterior cerebellar artery treated by neurosurgery and by endovascular embolization (Figure 4A, C, D). Shortly after, she also developed a right posterior communicating artery ICA treated by endovascular embolization and a small asymptomatic aneurysm on the right middle cerebral artery (Figure 4B). She later died from a hemorrhagic stroke. In addition, proband 2 was also diagnosed with hearing impairment, requiring a hearing aid device a few years before her death. Otoscopic examination of the ear was normal, but the audiometry tests revealed bilateral perceptive hearing impairment in the upper frequency range (Figure 4E, F). Two of the proband’s siblings were homozygous for the *SLC44A2* deletion and are phenotyped as VER–, and one of them (IV.4) developed an anti-VER alloantibody after RBC transfusion. Clinically, both siblings developed epileptic seizures and hearing loss from their thirties. The VER+ siblings with a heterozygous deletion of *SLC44A2* only suffer from hearing loss, while the wild-type VER+ sibling (IV.7) is healthy. In accordance with *Slc44a2*^−/–^ and *Slc44a2*^+/–^mice, the *SLC44A2* null allele in proband 2 and her siblings is associated with progressive age-related hearing impairment ^28^. Interestingly, in the proband 2 family, the homozygous *SLC44A2* null allele was associated with more severe diseases, including ICA and epilepsy.

**Figure 4:**
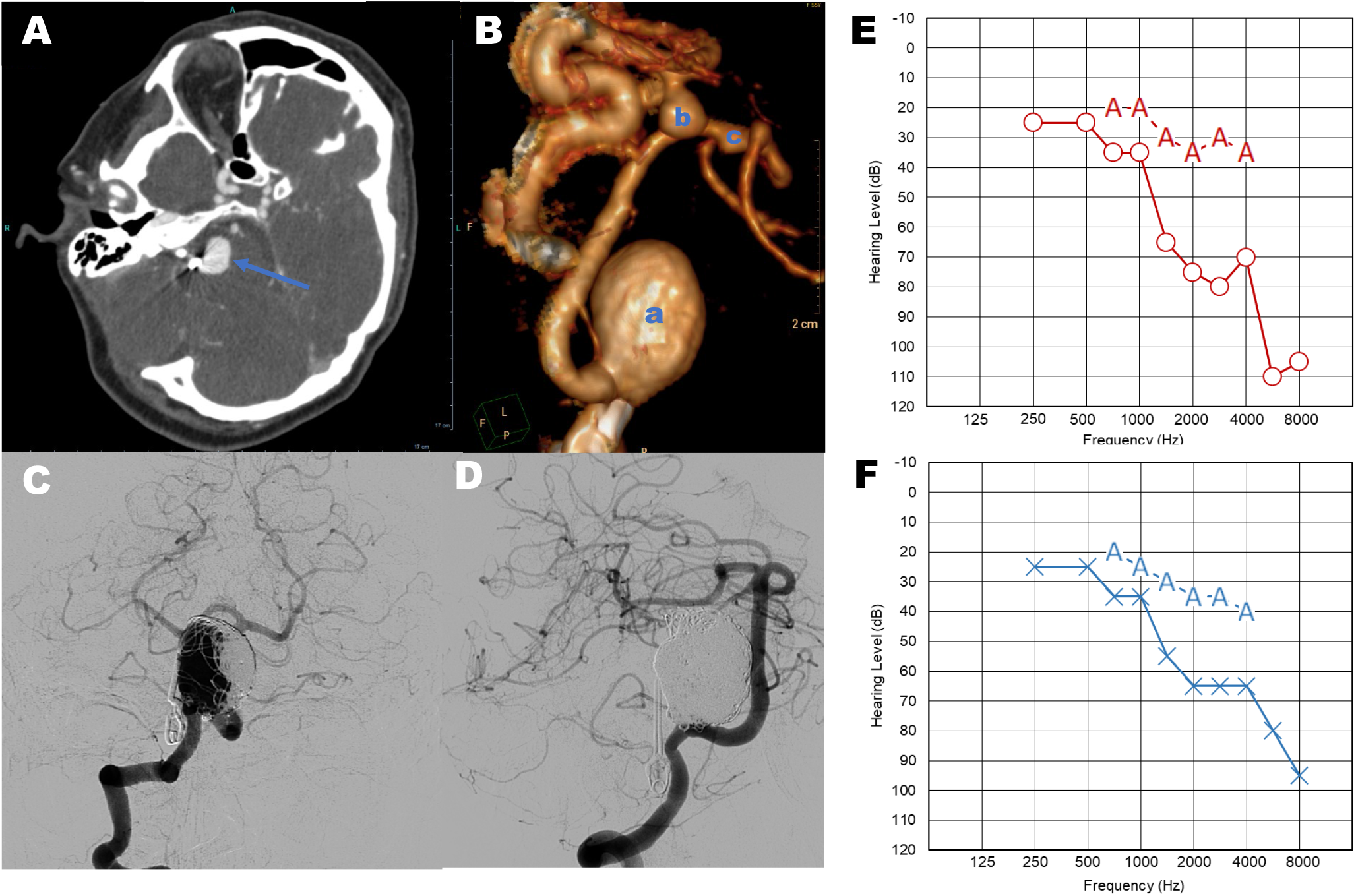
Clinical features of the VER– proband. **A**. CT angiography of the giant unruptured intracranial aneurysm on the right posterior cerebellar artery (PCA, blue arrow). **B**. 3D volume rendered of multiple cerebral aneurysms on the right PCA (a), on right posterior communicating artery (b), and on right middle cerebral artery (c). **C** and **D**. Angiograms before and after embolization of the PCA aneurysm, respectively. **E. and F**. Audiograms of VER-proband, showing a hearing loss by air conduction audiometry (blue line) and bone conduction audiometry (red line) for both left and right ear, witnessing a perception hearing loss in this patient.

### SLC44A2 is dispensable for erythropoiesis and platelet aggregation

Choline is an essential metabolite for cells because it plays an important role in the synthesis of the phospholipids that maintain membrane integrity. SLC44A2 was originally called CTL2 for choline transporter-like protein 2 because it has homology to other choline transporters. It belongs to the same SLC family as CTL1 (SLC44A1), which was discovered to complement a choline transport deficient yeast strain using RNA from the electric organ of the Torpedo fish^29,30^. SLC44A2 in the cell membrane was found to be a poor choline transporter when compared to its homolog SLC44A1, and its function is still not well understood^26^. To better appreciate the function of SLC44A2 in erythroid cells, we analyzed the rheological properties of the RBCs from the SLC44A2_null_ proband 2. The proband 2 RBCs displayed normal RBC deformability (Supplemental Figure 3), suggesting that the absence of SLC44A2 does not alter the biophysical properties of RBC membranes. Next, we compared the proliferation and differentiation of hematopoietic stem and progenitor cells (HSPCs) isolated from the peripheral blood of SLC44A2_null_ proband and from a healthy donor as a control. Markers used to assess the progression of erythroid differentiation included glycophorin A (GPA), band 3 and CD49d (α4 integrin) and showed normal *in vitro* erythroid proliferation and differentiation (Figure 5A and B), which is consistent with the absence of erythroid disorders in this proband and her VER– sibling (Supplemental Table 1). In addition, the monitoring of SLC44A2 expression during erythropoiesis from control HSCs using anti-VER antibody showed a higher expression level in immature erythroblasts (pro-erythroblast stage, day 2) followed by a progressive strong decrease upon erythroid terminal maturation (Figure 5C). On the other hand, a recent study in *Slc44a2*^−/–^ mice showed that Slc44a2 controls platelet activation via its function as a mitochondrial choline transporter^13^. Both CTL2_null_ patients (IV.1 and IV.5) showed normal complete blood count parameters, compared with reference values, including platelet count (Supplemental Table 1). To evaluate the role of SLC44A2 in human platelet activation, we assessed several *ex vivo* aggregation tests of platelets from one CTL2_null_ patient and one healthy donor in the presence of several agonists. In contrast to *Slc44a2*^−/–^ mice, the absence of SLC44A2 from human platelets did not alter platelet aggregation (Supplemental Table 2). It is noteworthy that the proband 2, as well as the 2 others homozygous SLC44A2_null_ probands, never exhibited any coagulation disorder like frequent bleeding (epistaxis, profuse menstruation, gingivorrhagia, etc…). In summary, we showed that human SLC44A2 is dispensable for *ex vivo* erythropoiesis and platelet aggregation.

**Figure 5:**
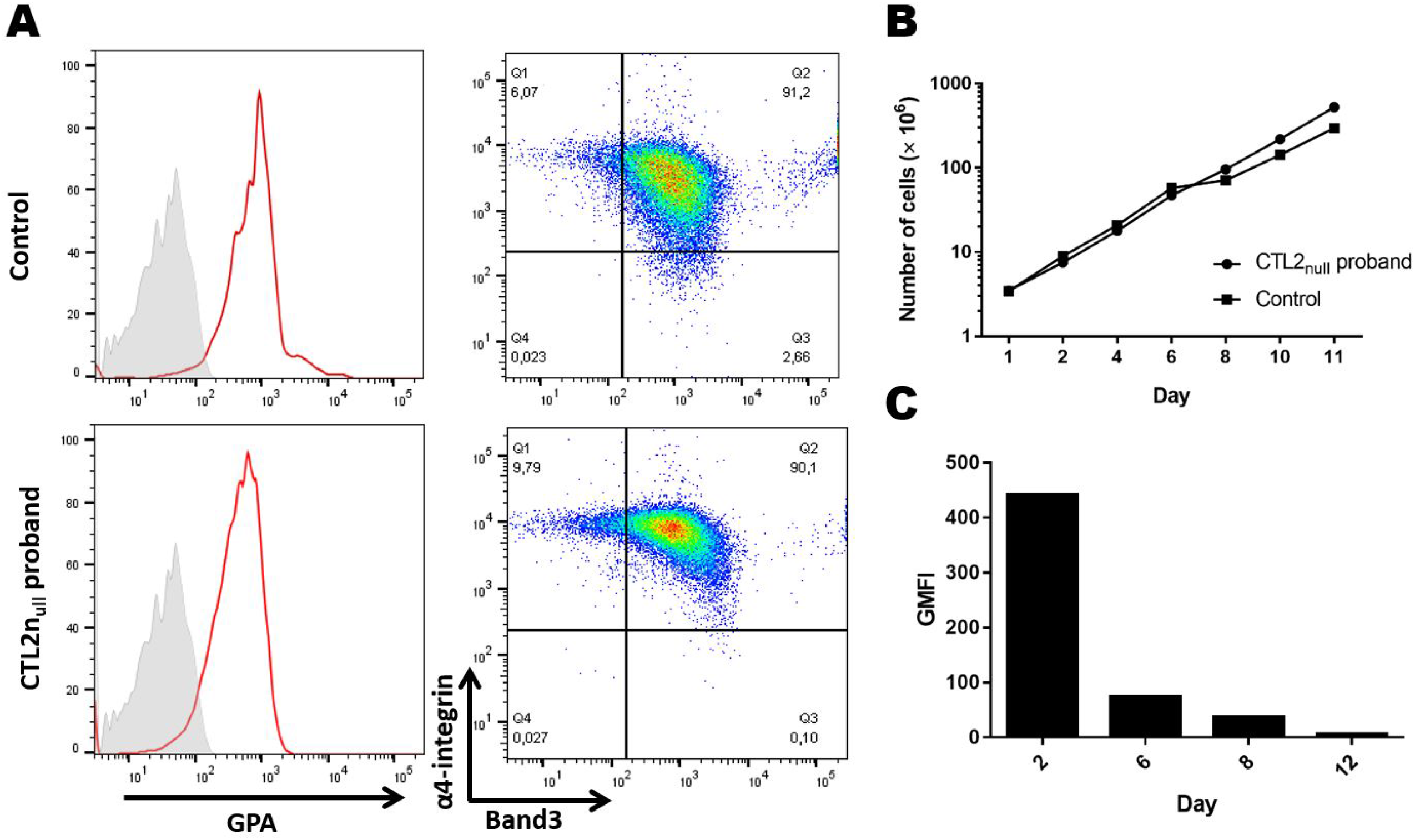
Normal *ex vivo* erythropoiesis of SLC44A2_null_ HSPCs. **A**. Terminal differentiation of HSPCs from healthy control and SLC44A2_null_ proband was monitored via α4-integrin and band 3 levels of GPA^pos^ cells at day 6 after EPO addition. **B**. Cell proliferation during *in vitro* erythropoiesis. **C**. CTL2 expression during terminal erythroid differentiation of HSPCs from healthy controls using an anti-VER antibody.

### Anti-SLC44A2 alloantibodies activate neutrophil adhesion to endothelial cells

SLC44A2 protein is highly expressed on neutrophils and carries the human neutrophil antigen-3 (HNA-3). Given that SLC44A2 is expressed in two isoforms (P1 and P2 transcripts), we compared the SLC44A2-specific cDNA derived from neutrophils and reticulocytes. We showed by RT-PCR using specific primers for both isoforms that reticulocytes expressed only the P1 variant, like neutrophils (Figure 6A). The absence of P1 and P2 transcripts in the PBMCs from the VER– proband 2 confirmed her null phenotype. Anti-HNA-3 antibodies were reported to be frequently involved in transfusion-related acute lung injury (TRALI) through neutrophil activation and aggregation in the pulmonary microvasculature. The HNA-3a/3b polymorphism results from a single nucleotide substitution (c.455G>A; p.Arg152Gln) in the first loop of CTL2 (Figure 6B). We next tested whether anti-VER and anti-RIF antibodies were reactive on neutrophils. Flow cytometry data showed that both antibodies recognized SLC44A2 protein in neutrophils (Figure 6C). To investigate the effect of these antibodies on neutrophil adhesion, neutrophils from healthy donors were incubated with both antibodies and then perfused through channels coated with endothelial cells, HMEC-1 (n=3). Both anti-VER and anti-RIF alloantibodies significantly increase neutrophil adhesion to endothelial cells compared to non-immune human IgG1 (*P* < .0001 and *P* < .001, respectively). This result suggested that anti-VER and anti-RIF antibodies may induce aggregates of neutrophils, such as anti-HNA-3 antibodies.

**Figure 6:**
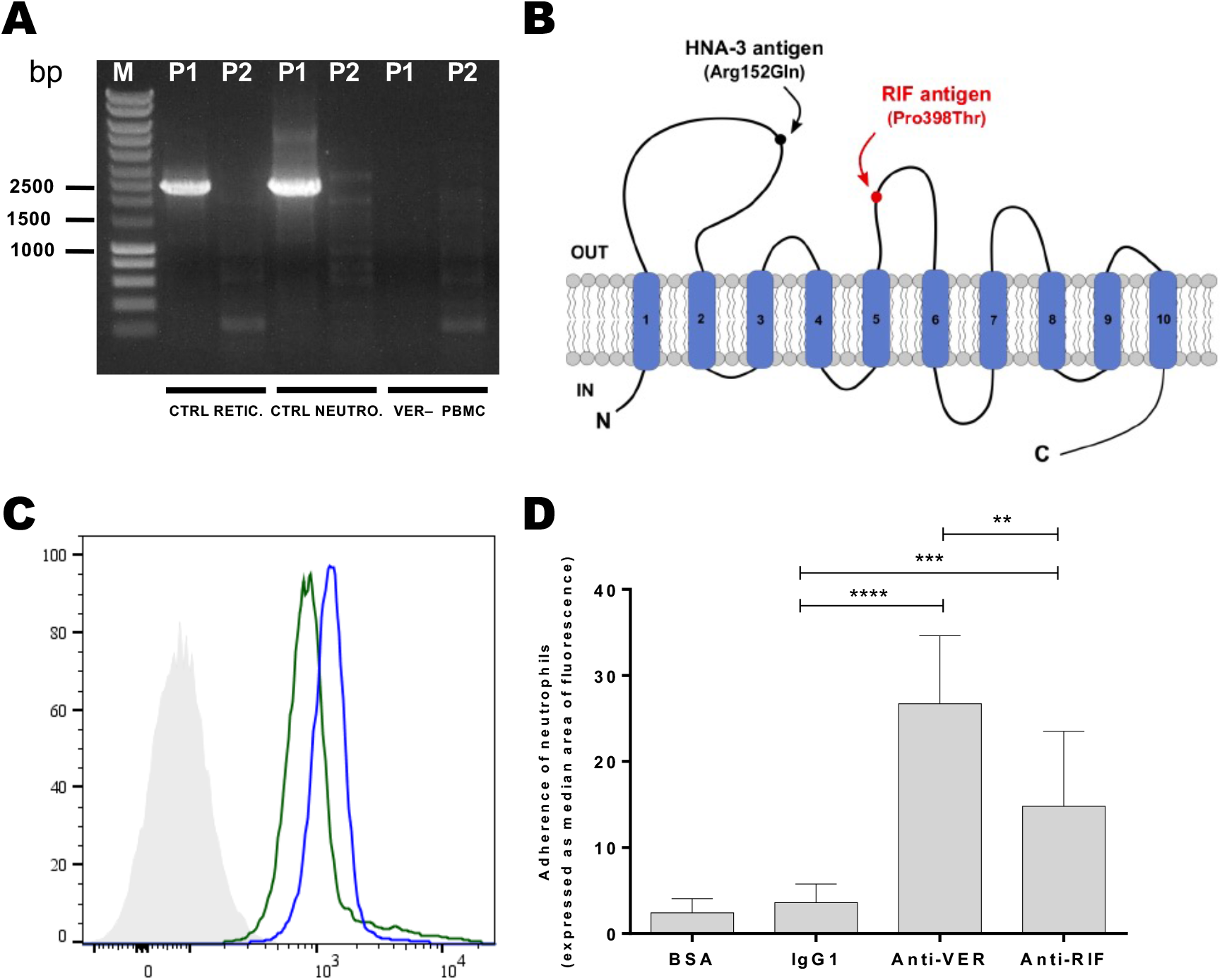
Anti-SLC44A2 red cell alloantibodies activate neutrophil adhesion to endothelial cells. **A**. SLC44A2 transcript variants P1 and P2 were amplified from cDNA samples isolated from control reticulocytes (CTRL RETIC), neutrophils (CTRL NEUTRO), and PBMC of the VER– proband 2 (VER PBMC). The purity of reticulocytes and neutrophils was verified by flow cytometry using anti-CD71 and anti-CD16 antibodies, respectively. **B**. Predicted topology of the human CTL2 choline transporter-like protein (SLC44A2, isoform P1) highlighting the location of the HNA-3 (Arg152Gln) and RIF (Pro398Thr) antigens; transmembrane domains are numbered 1 to 10. The P1 transcript variant encodes the shorter isoform of 704 amino acids in which arginine determining the HNA-3 antigen is at position 152. **C**. Flow cytometry analysis of neutrophil cells from control donors with anti-RIF (blue) and anti-VER (green) antibodies. The gray profile corresponds to neutrophils incubated with the secondary antibody only. **D**. Effect of anti-VER and anti-RIF on the adhesion of neutrophils from healthy controls to endothelial cells (HMEC-1). Adherent neutrophils were quantified and means ± standard error of the mean (SEM) of 3 independent experiments are shown. \*\**P* < .01, \*\*\**P* < .001, \*\*\*\**P* < .0001.

## Discussion

Here, we provide convincing evidence for the identification of a novel blood group system, as defined by two human RBC alloantibodies (anti-VER and anti-RIF), encoded by the *SLC44A2* gene located on chromosome 19p13.2. The gene product is the CTL2 protein (choline transporter-like protein 2), which carries two antigens as of today, the RIF and VER high-prevalence antigens. There was no history of hemolytic disease of the fetus and newborn, and acute hemolytic transfusion reaction in RIF– and VER– families, indicating that both antibodies are not clinically significant in RBC transfusion. However, these antibodies induce *ex vivo* neutrophil adhesion to endothelial cells, suggesting a potential implication within TRALI syndrome after plasma transfusion. In addition, our data confirm that CTL2 is expressed in human erythroid precursors and mature RBCs, but at a lower level compared to its expression on neutrophils (Figure 2B and 6C). The p.Pro398Thr variant found in RIF– North African individuals is predicted to be nonpathogenic by the SIFT and PolyPhen software and does indeed not significantly affect the level expression of SLC44A2 on RBCs (Figure 2B). However, the absence of erythroid defects in proband 2 and her sibling (IV.5), as well as normal *in vitro* erythropoiesis, indicate that SLC44A2 is dispensable for erythroid proliferation and differentiation. In neutrophils, CTL2 is highly expressed (∼ 5×10^5^ copies/cell)^31^. However, we have no evidence that the CTL2_null_ phenotype is associated with innate immunity dysfunction, as SLC44A2_null_ individuals have no significant history of severe infection. Rather, its absence in the two SLC44A2_null_ probands has no consequence on the neutrophil count in accordance with the *slc44a2*^*-/-*^ mice model (Supplemental Table 1), suggesting that other choline transporter(s) may compensate for the absence of SLC44A2 in these cells. Considering platelet function, a recent study in mice showed that Slc44a2 (KO) platelets aggregate less than Slc44a2 (WT) platelets after thrombin treatment^13^. Unexpectedly, human SLC44A2_null_ platelets displayed normal aggregation with several agonists including TRAP at different concentrations. These results are consistent with the absence of thromboembolic complication in the three SLC44A2_null_ individuals of this study. Collectively, the absence of hematological disorders in SLC44A2_null_ patients is particularly intriguing considering the reported function of this protein in mice model. An attractive hypothesis to explain this difference may be related to a possible genetic compensation in SLC44A2_null_ patients.

Nevertheless, *SLC44A2* polymorphisms or mutations in humans are far from harmless. Indeed, *SLC44A2* mutations have been identified in genome-wide association studies to have an allele-specific association with venous thromboembolism and cardiovascular diseases^15,16^. More recently, venous thromboembolism recurrence in Thai patients was shown to be linked to SLC44A2 p.Arg152Gln variant^32^. Considering our SLC44A2_null_ patient (proband 2), she suffered from multiple atypical ICAs, which characteristics (onset at a young age, large size, multiple sites, recurrence after treatment) are consistent with a congenital origin rather than a sporadic phenomenon^33-35^. In addition, the two other homozygous SLC44A2_null_ siblings were reported as having both epileptic disease occurring between 30 and 40 years old, but we lack the cerebral investigations allowing us to determine the presence of ICA in these patients. Nevertheless, it is difficult to assess the significance of the association between SLC44A2_null_ phenotype and these pathologies based only on this family case. Further investigations in additional unrelated SLC44A2_null_ individuals are necessary to fully appreciate the relevance of this observation. However, the progressive hearing impairment related to the *SLC44A2* deletion in this family is consistent with the hearing deficiency observed in the *slc44a2*^−/–^ mice, which was shown to be secondary to loss of auditory sensory cells, and spiral ganglion cells^28^. Our data strongly validate the crucial function of SLC44A2 in hearing maintenance.

In summary, our data demonstrate that defective alleles of *SLC44A2* occur in humans, do not preclude viability and are not necessarily associated with hematological disorders observed in murine models. However, the precise role of SLC44A2 in cerebrovascular maintenance remains to be established in mice and humans, especially as this protein is highly expressed in the brain-blood barrier.

## Materials & methods

### Subjects

Samples from subjects lacking a high-prevalence antigen of unknown specificity were cryopreserved at the National Reference Center for Blood Groups (CNRGS, biocollection #DC-2016-2872). Informed consent for research studies was obtained for all subjects in accordance with the Declaration of Helsinki protocols, and the study was approved by local institutional review boards.

### Clinical reports of the probands

Proband 1, a North African woman, was identified during her second pregnancy through a routine RBC antibody screen. Her serum was found to react 2+ with all RBCs tested, including a comprehensive panel of RBCs with a rare phenotype from the collections of the CNRGS, with the exception of her own RBCs. The antibody titration test was consistent with a so-called “HTLA” (high titer low affinity) profile (titer 512, 2+ reactivity until dilution 1/256). At 37 weeks of gestation, the proband naturally gave birth to a healthy boy. We decided to name this antibody anti-RIF.

Proband 2, a European woman was hospitalized for intraparenchymal hemorrhage due to a fall. Her clinical history showed a giant cerebral aneurysm at the posterior inferior cerebellar region, treated by surgical clipping, followed by a plausible transfusion. Her serum was positive with a large panel of RBCs with a rare phenotype (negative autocontrols), with no possibility of finding an antibody specificity. We decided to name this antibody anti-VER after the proband’s city of birth.

### Blood group serology and subjects screening

Antibody identification and RBC typing were performed by indirect antiglobulin gel-test (ID-Card LISS/Coombs; DiaMed, Bio-Rad). The serum of proband 1 containing anti-RIF was used to screen compatible subjects from cryopreserved RBCs lacking a high-prevalence antigen of unknown specificity.

### Exome sequencing and data analysis

Genomic DNA was extracted from whole blood cells by using a benchtop instrument for fully automated nucleic acid purification (MagNaPure Compact, Roche Life Science) according to the manufacturer’s recommendations. Exome capture was performed with the Sure Select Human All Exon Kit (Agilent Technologies). Agilent Sure Select Human All Exon (58 Mb, V6) libraries were prepared from 3 µg of genomic DNA sheared with an Ultrasonicator (Covaris) as recommended by the manufacturer. Barcoded exome libraries were pooled and sequenced with a HiSeq2500 system (Illumina), generating paired-end reads. After demultiplexing, sequences were mapped on the human genome reference (NCBI build 37, hg19 version) with BWA. The mean depth of coverage obtained for RIF– individuals exome libraries was >120X with >=96% and >=94% of the targeted exonic bases covered at least 15 and 30 independent sequencing reads. Variant calling was carried out with the Genome Analysis Toolkit (GATK), SAMtools, and Picard tools. Single nucleotide variants were called with GATK Unified Genotyper, whereas indel calls were made with the GATK IndelGenotyper_v2.

All variants with read coverage 23% and Phred-scaled quality 20% were filtered out with PolyWeb, an in-house-developed annotation software.

### Sanger sequencing

Exon 14 of the *SLC44A2* gene was amplified with primers SLC44A2-P1F (forward) and SLC44A2-P2R (reverse) from genomic DNA (Supplemental Table 3). The PCR product was sequenced using the SLC44A2-S1F primer. The region containing the deletion breakpoints in the *SLC44A2* gene of the VER– proband 2 were amplified with PCR primers SLC44A2-P3F and SLC44A2-P4R and sequenced using SLC44A2-P2R. Detailed PCR conditions are available upon request.

### Genotyping

Genotyping of the RIF– proband 1 family was performed in genomic DNA by allele-specific PCR (AS-PCR) analysis. Two PCRs were performed with specific reverse primers: SLC44-1192C-R for the 1192C allele, SLC44-1192A-R for the 1192A allele and a common forward primer SLC44A2-P5F. The PCR products were interpreted on the basis of positive or negative amplification. As an internal control, two additional primers (hGH-F and hGH-R) for *hGH* amplification were used (Supplemental Table 3).

### Erythroid cell culture and characterization

Peripheral blood mononuclear cells (PBMCs) were obtained from blood samples of healthy donors and VER– proband 2 using Ficoll density gradient separation (Pancoll 1.077 g/ml, Pan-Biotech). CD34+ cells were purified using anti-CD34 conjugated microbeads and manual cell separation columns according to the manufacturer’s instructions (Miltenyi). The cell culture procedure was comprised of 2 phases as described by Freyssinier *et al*^*36*^. CTL2 expression during terminal erythroid differentiation of HSCs from healthy controls was monitored by flow cytometry using an anti-VER antibody.

### Plasmid construction

The pcDNA3.1/V5-HisTOPO plasmid containing the full-length CTL2 cDNA (NM_001145056.1) was kindly provided by Sentot Santoso^37^. Plasmids with the c.1192A mutation were prepared using the QuikChange XL Site-Directed Mutagenesis Kit (Stratagene) according to the manufacturer’s protocol. A detailed mutagenesis PCR protocol is available upon request.

### Cell culture and transfection

L-929 cells were grown in DMEM (Gibco) supplemented with 10% fetal bovine serum (Pan-Biotech) and 0.5X antibiotic-antimycotic solution (Gibco) at 37°C under a humidified atmosphere containing 5% CO_2_. Then, 3.10^5^ L-929 cells were transfected with 1 µg of the plasmid using FuGENE® 6 Transfection Reagent as recommended by the manufacturer (Roche). Stable L-929 transfectants were obtained after three weeks of selection with Geneticin (1 mg/mL, Thermo Fisher Scientific).

### Flow cytometry analyses in RBCs and L-929 cells

Anti-RIF and anti-VER antibodies were purified by adsorption-elution of sera from the RIF– and VER– probands, respectively, on papain-treated RBCs using the Gamma Elu Kit II (Immucor). Thawed RBCs or fresh L-929 cells were washed in Dulbecco’s phosphate-buffered saline solution (Gibco), resuspended in low-ionic strength BFI buffer supplemented with 1% BSA and incubated with purified antibodies (1:2) at 37°C for one hour. Anti-RIF and anti-VER labeling were revealed with goat F(ab’)2 anti-human IgG(H + L)-PE (1:50; Beckman Coulter) and immediately analyzed with a FACSCantoII flow cytometer (BD Bioscience).

### Immunoprecipitation, Sodium Dodecyl Sulfate Polyacrylamide Gel Electrophoresis, and Western Blotting

Recombinant human SLC44A2 was produced in Sf9 insect cells transfected with full-length *SLC44A2* cDNA as described^25^. SLC44A2–NT (CTL2-NT) rabbit antiserum against synthetic SLC44A2 peptides was coupled to CNBr beads as previously described and used to immunoprecipitate SLC44A2 from cell lysates as described^27^. Whole cell lysates from lung tissues of Slc44a2 wild-type and knockout FVB mice^28^ were prepared in lysis buffer with 1% NP-40 and stored frozen until use. UM-SCC-47, a human squamous cell carcinoma cell line, naturally expresses wild-type SLC44A2 and served as a source of wild-type SLC44A2 protein. Cell lysates of Sf9 insect cells expressing full-length rHuSLC44A2 and UM-SCC 47 expressing wild-type SLC44A2 protein were similarly prepared. The cell lysates (150 µg protein) were immunoprecipitated with anti CTL2-NT beads at a concentration of 2 µg/ml, collected and washed by centrifugation and subjected to sodium dodecyl sulfate polyacrylamide gel electrophoresis (SDS-PAGE) as described previously^25^. The primary antibodies, rabbit anti-CTL2-NT (1/500 or 2 µg/ml), human alloantibodies RIF and VER were used at 1:10. Antibody binding on Western blots was detected with enhanced chemiluminescence using the appropriate affinity purified secondary antibody. Rabbit antihuman IgG-IGM-specific antiserum was diluted 1:2000. Goat anti-rabbit IgG heavy and light chain specific was used at 1:5000.

### Neutrophil and reticulocyte isolation

Human neutrophils were isolated from fresh whole blood (< 4 hours after blood sampling) using the MACSxpress Neutrophil Isolation Kit followed by a MACSxpress Erythrocyte Depletion Kit (Miltenyi Biotec). Reticulocytes were isolated from whole blood using the CD71 MicroBeads Kit (Miltenyi Biotec). The purity of the cells, determined by flow cytometry using anti-CD16 and anti-CD71 antibodies, was approximately 98%.

### Amplification of *SLC44A2* transcripts

mRNA was extracted from neutrophils and reticulocytes and transcribed with superScript III first-strand synthesis SuperMix, according to the manufacturer’s instructions (Thermo Fisher Scientific). cDNA products were amplified by PCR using a primer combination specific for the P1 and P2 CTL2 transcripts. Primer sequences are available in Supplemental Table 3.

### Rheology experiments

RBC deformability (EI, elongation index) was determined at 37°C at 9 shear stresses ranging from 0.30 to 30 Pa by laser diffraction analysis (ektacytometry) using the laser-assisted optical rotational cell analyzer (LORCA, RR Mechatronics).

### Platelet aggregation studies

Platelet aggregation tests in the presence of several agonists were measured with an optical platelet aggregometer (APACT 4004) according to the manufacturer’s instructions.

### Flow neutrophil adhesion assays

HMEC-1 (Human Microvascular Endothelial Cell line) monolayers were grown in Vena8 Endothelial+ Biochips (Cellix Ltd, Dublin, Ireland) as previously described^38^, and activated by TNFα (10 ng/ml) for 24 h prior to the adhesion assay. In brief, adhesion was measured under flow conditions using Vena8 Endothelial+TM biochips (internal channel dimensions: length 20 mm, width 0.8 mm, height 0.12 mm) and ExiGoTM Nanopumps (Cellix Ltd, Dublin, Ireland). Neutrophils were incubated with anti-RIF, anti-VER or irrelevant IgG1 antibodies 30 min before cell perfusion. Samples were then perfused for 45 min at 1 dyn/cm2 through the biochip channels containing HMEC-1 monolayers. Adherent neutrophils were labeled with anti-CD16 alexa 488-conjugated mouse monoclonal antibody (Biolegend, San Diego, USA) for 15 min. Neutrophil adhesion was monitored using AxioObserver Z1 microscope and ZEN software (Carl Zeiss, Le Pecq, France). Images were taken in 11 representative fields at the centerline of each channel at 10 min intervals throughout the assay. Adhesion levels were quantified in the last images set, by measuring the surface area of fluorescent patches using ImageJ Software. Adhesion in each channel is defined as the median value of fluorescence of the 11 fields.

## Data Availability

All data produced in the present work are contained in the manuscript

## Acknowledgments

We thank the patients and their families for their participation in the study. We thank all members of Centre National de Référence pour les Groupes Sanguins (CNRGS) for the management of blood samples and immunohematological studies. We thank Laurent Chabre (Oriade Noviale laboratory, Voreppe, France) for his help in organizing blood sample collection. We are indebted to William Vainchenker for helpful comments and for reading the manuscript. We are obliged to Ghania Hariti and Soraya Iguergaziz for helpful comments. This work was supported by a grant from l’Association Recherche et Transfusion (#151/2016), the Institut National de la Transfusion Sanguine, and the Laboratory of Excellence GR-Ex, reference ANR-11-LABX-0051; GR-Ex is funded by the program “Investissements d’avenir” of the French National Research Agency, reference ANR-11-IDEX-0005-02.

## Authorship Contributions

J.P.C., O.H., C.L.V.K., Y.C., S.A. and T.P. conceived and designed the study. C.V., B.K., M.M., T.S.N., L.Y., and G.L. performed the experiments. C.V., B.K., M.M., T.E.C., S.A., and T.P. analyzed the data. S.A.M., C.G.L., H.E.K. and O.D. contributed biologic materials and collected the clinical history of RIF– and VER– individuals. S.A wrote the paper. J.P.C., C.L.V.K., N.M., T.EC. and T.P. reviewed the manuscript.

## References

1. Storry JR, Clausen FB, Castilho L, et al. International Society of Blood Transfusion Working Party on Red Cell Immunogenetics and Blood Group Terminology: Report of the Dubai, Copenhagen and Toronto meetings. Vox Sang. 2019;114(1):95–102.

2. Saison C, Helias V, Ballif BA, et al. Null alleles of ABCG2 encoding the breast cancer resistance protein define the new blood group system Junior. Nat Genet. 2012;44(2):174–177.

3. Helias V, Saison C, Ballif BA, et al. ABCB6 is dispensable for erythropoiesis and specifies the new blood group system Langereis. Nat Genet. 2012;44(2):170–173.

4. Mikdar M, Gonzalez-Menendez P, Cai X, et al. The equilibrative nucleoside transporter ENT1 is critical for nucleotide homeostasis and optimal erythropoiesis. Blood. 2021;137(25):3548–3562.

5. Azouzi S, Mikdar M, Hermand P, et al. Lack of the multidrug transporter MRP4/ABCC4 defines the PEL-negative blood group and impairs platelet aggregation. Blood. 2020;135(6):441–448.

6. Duval R, Nicolas G, Willemetz A, et al. Inherited glycosylphosphatidylinositol defects cause the rare Emm-negative blood phenotype and developmental disorders. Blood. 2021;137(26):3660–3669.

7. Daniels G, Ballif BA, Helias V, et al. Lack of the nucleoside transporter ENT1 results in the Augustine-null blood type and ectopic mineralization. Blood. 2015;125(23):3651–3654.

8. Bertelson CJ, Pogo AO, Chaudhuri A, et al. Localization of the McLeod locus (XK) within Xp21 by deletion analysis. Am J Hum Genet. 1988;42(5):703–711.

9. Ribeiro ML, Alloisio N, Almeida H, et al. Severe hereditary spherocytosis and distal renal tubular acidosis associated with the total absence of band 3. Blood. 2000;96(4):1602–1604.

10. Cherif-Zahar B, Raynal V, Gane P, et al. Candidate gene acting as a suppressor of the RH locus in most cases of Rh-deficiency. Nat Genet. 1996;12(2):168–173.

11. Preston GM, Smith BL, Zeidel ML, Moulds JJ, Agre P. Mutations in aquaporin-1 in phenotypically normal humans without functional CHIP water channels. Science. 1994;265(5178):1585–1587.

12. Azouzi S, Gueroult M, Ripoche P, et al. Energetic and molecular water permeation mechanisms of the human red blood cell urea transporter B. PLoS One. 2013;8(12):e82338.

13. Bennett JA, Mastrangelo MA, Ture SK, et al. The choline transporter Slc44a2 controls platelet activation and thrombosis by regulating mitochondrial function. Nat Commun. 2020;11(1):3479.

14. Zirka G, Robert P, Tilburg J, et al. Impaired adhesion of neutrophils expressing Slc44a2/HNA-3b to VWF protects against NETosis under venous shear rates. Blood. 2021;137(16):2256–2266.

15. Germain M, Chasman DI, de Haan H, et al. Meta-analysis of 65,734 individuals identifies TSPAN15 and SLC44A2 as two susceptibility loci for venous thromboembolism. Am J Hum Genet. 2015;96(4):532–542.

16. Hinds DA, Buil A, Ziemek D, et al. Genome-wide association analysis of self-reported events in 6135 individuals and 252 827 controls identifies 8 loci associated with thrombosis. Hum Mol Genet. 2016;25(9):1867–1874.

17. Tilburg J, Coenen DM, Zirka G, et al. SLC44A2 deficient mice have a reduced response in stenosis but not in hypercoagulability driven venous thrombosis. J Thromb Haemost. 2020;18(7):1714–1727.

18. Constantinescu-Bercu A, Grassi L, Frontini M, Salles C, II, Woollard K, Crawley JT. Activated alphaIIbbeta3 on platelets mediates flow-dependent NETosis via SLC44A2. Elife. 2020;9.

19. Inazu M. Functional Expression of Choline Transporters in the Blood-Brain Barrier. Nutrients. 2019;11(10).

20. Iwao B, Yara M, Hara N, et al. Functional expression of choline transporter like-protein (CTL1) and CTL2 in human brain microvascular endothelial cells. Neurochem Int. 2016;93:40–50.

21. Greinacher A, Wesche J, Hammer E, et al. Characterization of the human neutrophil alloantigen-3a. Nat Med. 2010;16(1):45–48.

22. Flesch BK, Wesche J, Berthold T, et al. Expression of the CTL2 transcript variants in human peripheral blood cells and human tissues. Transfusion. 2013;53(12):3217–3223.

23. Bryk AH, Wisniewski JR. Quantitative Analysis of Human Red Blood Cell Proteome. J Proteome Res. 2017;16(8):2752–2761.

24. Tilburg J, Adili R, Nair TS, et al. Characterization of hemostasis in mice lacking the novel thrombosis susceptibility gene Slc44a2. Thrombosis research. 2018;171:155–159.

25. Kommareddi PK, Nair TS, Vallurupalli M, et al. Autoantibodies to recombinant human CTL2 in autoimmune hearing loss. Laryngoscope. 2009;119(5):924–932.

26. Kommareddi PK, Nair TS, Thang LV, et al. Isoforms, expression, glycosylation, and tissue distribution of CTL2/SLC44A2. Protein J. 2010;29(6):417–426.

27. Nair TS, Kozma KE, Hoefling NL, et al. Identification and characterization of choline transporter-like protein 2, an inner ear glycoprotein of 68 and 72 kDa that is the target of antibody-induced hearing loss. J Neurosci. 2004;24(7):1772–1779.

28. Kommareddi P, Nair T, Kakaraparthi BN, et al. Hair Cell Loss, Spiral Ganglion Degeneration, and Progressive Sensorineural Hearing Loss in Mice with Targeted Deletion of Slc44a2/Ctl2. J Assoc Res Otolaryngol. 2015;16(6):695–712.

29. O’Regan S, Traiffort E, Ruat M, Cha N, Compaore D, Meunier FM. An electric lobe suppressor for a yeast choline transport mutation belongs to a new family of transporter-like proteins. Proc Natl Acad Sci U S A. 2000;97(4):1835–1840.

30. O’Regan S, Meunier FM. Selection and characterization of the choline transport mutation suppressor from Torpedo electric lobe, CTL1. Neurochem Res. 2003;28(3-4):551–555.

31. Rieckmann JC, Geiger R, Hornburg D, et al. Social network architecture of human immune cells unveiled by quantitative proteomics. Nat Immunol. 2017;18(5):583–593.

32. Apipongrat D, Numbenjapon T, Prayoonwiwat W, Arnutti P, Nathalang O. Association between SLC44A2 rs2288904 polymorphism and risk of recurrent venous thromboembolism among Thai patients. Thrombosis research. 2019;174:163–165.

33. Caranci F, Briganti F, Cirillo L, Leonardi M, Muto M. Epidemiology and genetics of intracranial aneurysms. Eur J Radiol. 2013;82(10):1598–1605.

34. Krischek B, Inoue I. The genetics of intracranial aneurysms. J Hum Genet. 2006;51(7):587–594.

35. Alg VS, Sofat R, Houlden H, Werring DJ. Genetic risk factors for intracranial aneurysms: a meta-analysis in more than 116,000 individuals. Neurology. 2013;80(23):2154–2165.

36. Freyssinier JM, Lecoq-Lafon C, Amsellem S, et al. Purification, amplification and characterization of a population of human erythroid progenitors. Br J Haematol. 1999;106(4):912–922.

37. Bayat B, Tjahjono Y, Sydykov A, et al. Anti-human neutrophil antigen-3a induced transfusion-related acute lung injury in mice by direct disturbance of lung endothelial cells. Arterioscler Thromb Vasc Biol. 2013;33(11):2538–2548.

38. Koehl B, Nivoit P, El Nemer W, et al. The endothelin B receptor plays a crucial role in the adhesion of neutrophils to the endothelium in sickle cell disease. Haematologica. 2017;102(7):1161–1172.

