## Supplemental data for "Lack of the human choline transporter-like protein CTL2 causes hearing impairment and a rare red blood cell phenotype"

### SUPPLEMENTAL TABLES

#### Supplemental Table 1: Hematological parameters of RIF– and VER– probands.

| | RIF–probands ( p.Pro398Thr) | | | VER–probands ($CTL2_{null}$) | | |
|---|---|---|---|---|---|---|
| | **1** | **2 (proband 1)** | **3** | **IV.1 (proband 2)** | **IV.5** | **Reference values** |
| RBCs ($10^{12}$/L) | 3.8 | 4.8 | 4.2 | 4.6 | 4.7 | 4-5 |
| HGB (g/dL) | 11.4 | 14.3 | 13.6 | 13.3 | 15 | 11-15 |
| HCT (%) | 35 | 41.6 | 39.7 | 41.1 | 46.5 | 35-50 |
| MCV (fL) | 90.7 | 86.7 | 93.6 | 91.6 | 97.7 | 82-98 |
| MCH (pg) | 29.4 | 29.8 | 32.1 | 29 | 31.5 | 27-32 |
| MCHC (g/dL) | 32.4 | 34.4 | 34.3 | 33.1 | 32.3 | 32-35 |
| WBCs ($10^9$/L) | 7.3 | 6.5 | 4.4 | 7.9 | 7.6 | 4 -10 |
| Neutrophils ($10^9$/L) | N.D | N.D | N.D | 4.5 | N.D | 2-8 |
| Platelets ($10^9$/L) | 331 | 331 | 157 | 278 | 294 | 150-400 |
| Reticulocytes ($10^9$/L) | N.D | N.D | N.D | 28 | N.D | 20-150 |

#### Supplemental Table 2: Aggregation tests of platelets from healthy donor (control) and CTL2null patient (Proband 2).

| | Max. aggregation | | |
|---|---|---|---|
| | Index patient (%) | Index control (%) | Reference values (%) |
| Collagen (2 µg/mL) | 24 | 78 | >55 |
| Collagen (5 µg/mL) | 67 | 76 | >61 |
| ADP (2.5 mM) | 49 | 75 | >42 |
| ADP (5 mM) | 75 | 79 | >51 |
| Arachidonic acid (1mM) | 92 | 80 | >60 |
| Adrenaline (5 µM) | 84 | 77 | >65 |
| TRAP (10 µM) | 30 | 70 | 0-100 |
| TRAP (25 µM) | 81 | 81 | >71 |
| Ristocetin (1 mg/mL) | 87 | 89 | >60 |

**Supplementary Table 3 : Primers and PCR conditions used to confirm the c.1192C>A mutation by sequencing, the *SLC44A2* deletion breakpoint, the RIF genotyping and analysis of *SLCC4A2* P1 and P2 isoforms.**

| | **Assay** | **User Name** | **Sequence (5' to 3')** | **Location** | **Position** | **Product size** |
|---|---|---|---|---|---|---|
| c.1192C>A polymorphism confirmation | PCR | SLC44A2-P1F | CTGTGGGATACGTCATGTGC | Upstream of exon 12 | chr19 : 10635165 - 10635184 | 1221 bp |
| | | SLC44A2-P2R | GTAGAAGGCGAACTGGCAAC | Exon 15 | chr19 : 10636385 - 10636366 | |
|  | Sequencing | SLC44A2-S1F | AGTGACCTGCAGCTTAGGGA | Upstream of exon 13 | chr19 : 10635318 10635337 | - |
| SLC44A2 deletion breackpoint | PCR | SLC44A2-P3F | GGAGTTTCCCAGCCTACCTC | Upstream of 5'UTR | chr19 : 10598153 - 10598172 | 1331 bp |
| | | SLC44A2-P4R | AACCGTGGGTGGGACGTA | Downstream of exon 16 | chr19 : 10636775 - 10636758 | |
| | Sequencing | SLC44A2-P2R | GTAGAAGGCGAACTGGCAAC | Exon 15 | chr19 : 10636385 - 10636366 | - |
| RIF Genotyping | ASP-PCR | SLC44A2-P5F | TAAATGGGGGCAGTTGTAGC | Upstream of exon 11 | chr19 : 10634679 10634698 | 821 bp |
| | | SLC44-1192C-R<br>or<br>SLC44-1192A-R | TTCGCAGTAAATGGGCAGGG<br><br>TTCGCAGTAAATGGGCAGGT | On c.1192C>A polymorphism | chr19 : 10635499 - 10635480 | |
| | Amplification Control | hGH-F | TGCCTTCCCAACCATTCCCTTA | Exon 2 of *hGH* gene | chr17 : 63918421 - 63918442 | 434 bp |
|  |  | hGH-R | CCACTCACGGATTTCTGTTGTGTTTC | Exon 3 of *hGH* gene | chr17 : 63918009 63918034 |  |
| cDNA analysis | P1 Isoform PCR | SLC44A2-cP1F | ATGGAGGACGAGCGGAAA | Exon 1a | chr19 : 10602531 - 10602548 | 2291 pb |
| | | SLC44A2-3'UTR | TAAGAAGGAGATGCCCCAGA | 3' UTR | chr19 : 10643561 - 10643542 | |
| | P2 Isoform PCR | SLC44A2-cP2F | ATGGGGGACGAGCGGCCCCA | Exon 1b | chr19 : 10625634 - 10625653 | 2297 pb |
| | | SLC44A2-3'UTR | TAAGAAGGAGATGCCCCAGA | 3' UTR | chr19 : 10643561 - 10643542 | |

**SUPPLEMENTAL FIGURES**

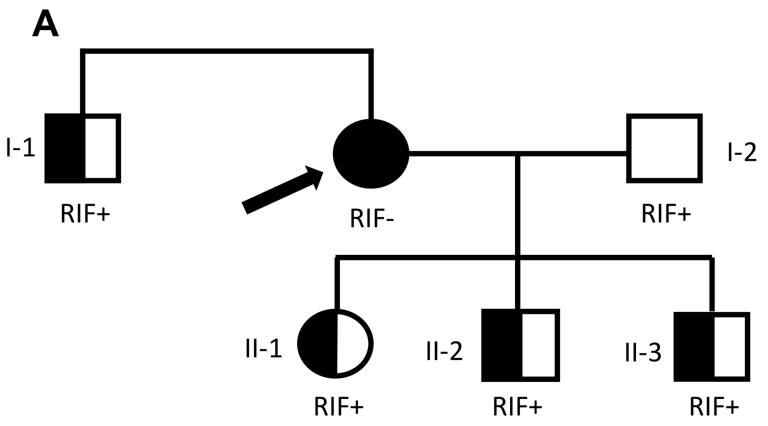


**Supplemental Figure 1**: Pedigree of the family of the RIF-proband indicating the genotype for each individual. Propositus is indicated by an arrow and presents the new mutation in homozygous state.

```
CTL2_HUMAN MEDERKNGAYGTPQKYDPTFKGPIYNRGCTDIICCVFLLLAIVGYVAVGIIAWTHGDPRK 60
CTL2_MOUSE MEDDRKDAVYGTPQKYDPTFKGPIYNRGCTDVICCVLLFLAIVGYVAVGIIAWTHGDPRK 60
           ***:**:..**********************:****:*:*********************

CTL2_HUMAN VIYPTDSRGEFCGQKGTKNENKPYLFYFNIVKCASPLVLLEFQCPTPQICVEKCPDRYLT 120
CTL2_MOUSE VIYPTDSRGEFCGQKGTKNADKPFLFYFNIVKCANPLVLLEFHCPTPQICVKQCPDRYLT 120
           ******************* :**:**********.*******:********::*******

CTL2_HUMAN YLNARSSRDFEYYKQFCVPGFKNNKGVAEVLQDGDCPAVLIPSKPLARRCFPAIHAYKGV 180
CTL2_MOUSE LLSARNTRDFDYYKQFCVPGFQNNKGVTEILRDGECPAVITPSKPLAQRCFPAIHASKGV 180
            *.**.:***:**********:*****:*:*:**:****: ******:******** ***

CTL2_HUMAN LMVGNETTYEDGHGSRKNITDLVEGAKKANGVLEARQLAMRIFEDYTVSWYWIIIGLVIA 240
CTL2_MOUSE LMVGNETTYEDGHGARKNITDLVEGAKKANKILEARQLAMQIFEDYTVSWYWIIIGLVIA 240
           **************:*************** :********:*******************

CTL2_HUMAN MAMSLLFIILLRFLAGIMVWVMIIMVILVLGYGIFHCYMEYSRLRGEAGSDVSLVDLGFQ 300
CTL2_MOUSE MVLSLLFIVLLRFLAGIMVWVMIVMVILVLGYGIFHCYMEYSRLRGEAGSDVSLVDLGFQ 300
           *.:*****:**************:************************************

CTL2_HUMAN TDFRVYLHLRQTWLAFMIILSILEVIIILLLIFLRKRILIAIALIKEASRAVGYVMCSLL 360
CTL2_MOUSE TDLRVYLHLRQTWMAFMIILSILEVVIILLLIFLRKRILIAIALIKEASRAVGHVMCSLL 360
           **:**********:***********:***************************:******

CTL2_HUMAN YPLVTFFLLCLCIAYWASTAVFLSTSNEAVYKIFDDSPCPFTAKTCNPETFPSSNESRQC 420
CTL2_MOUSE YPLVTFFLLCLCIAYWASTSVFLSTSNTAVYKVVDDTACPLLRKTCNPETFPLRNESLQC 420
           *******************:******* ****:.**: **:  *********  *** **

CTL2_HUMAN PNARCQFAFYGGESGYHRALLGLQIFNAFMFFWLANFVLALGQVTLAGAFASYYWALRKP 480
CTL2_MOUSE PTARCQFAFYGGESTYHRALLGLQIFNAFMFFWLANFVLALGQVTLAGAFASYYWAMRKP 480
           *.************ *****************************************:***

CTL2_HUMAN DDLPAFPLFSAFGRALRYHTGSLAFGALILAIVQIIRVILEYLDQRLKAAENKFAKCLMT 540
CTL2_MOUSE DDMPAFPLFSAFGRALRYHTGSLAFGSLILAIVQIIRVMLEYLDQRLKAAQNKFAKFLMV 540
           **:***********************:***********:***********:***** **.

CTL2_HUMAN CLKCCFWCLEKFIKFLNRNAYIMIAIYGTNFCTSARNAFFLLMRNIIRVAVLDKVTDFLF 600
CTL2_MOUSE CLKCCFWCLEKFIKFLNRNAYIMIAIYGTNFCTSARNAFFLLMRNIIRVAVLDKVTDFLF 600
           ************************************************************

CTL2_HUMAN LLGKLLIVGSVGILAFFFFTHRIRIVQDTAPPLNYYWVPILTVIVGSYLIAHGFFSVYGM 660
CTL2_MOUSE LLGKLLIVGSVGILAFFFFTHRIRIVQDTAPPLNYYWVPILTVIIGSYLIAHGFFSVYGM 660
           ********************************************:***************

CTL2_HUMAN CVDTLFLCFLEDLERNDGSAERPYFMSSTLKKLLNKTNKKAAES 704
CTL2_MOUSE CVDTLFLCFLEDLERNDGSAERPYFMSSTLKKLLNKTNKKVAES 704
           ****************************************.***
```

**Supplemental Figure 2**: Alignment of the primary amino acid sequences of human and mouse SLC44A2 (CTL2). The red box corresponds to the amino acid (398Pro) involved in the RIF+/RIF- polymorphism.

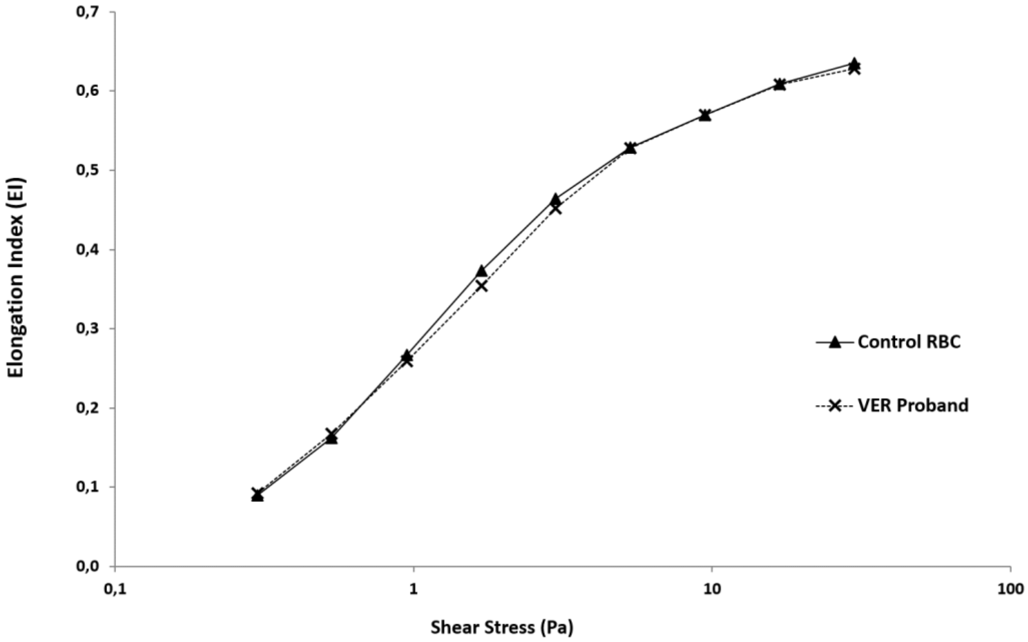


**Supplementary Figure 3:** The elongation index (i.e., deformability) of red blood cells (RBC) from VER proband and healthy control determined at discrete shear stresses between 0.5 and 30.0 Pa.
